# Malnutrition Diagnosis: Validation of a Weight-Independent “GLIM-Adapted” Framework in U.S. Adults (NHANES 1999–2020)

**DOI:** 10.64898/2026.09.04.26362229

**Authors:** F. Orsini, A. Novella, L. Pasina

## Abstract

The Global Leadership Initiative on Malnutrition (GLIM) consensus standardized malnutrition diagnosis, but incorporating low Body Mass Index (BMI) as a phenotypic criterion introduces a “circularity bias” in weight-stratified analyses and conceals “hidden hunger” in high-obesity populations. Using nationally representative data from the U.S. NHANES 1999–2020 (N = 23,363 adults), we propose a “GLIM-adapted” framework that excludes BMI—relying solely on weight loss and DXA-derived appendicular lean mass index (ALMI). Standard GLIM identified an 18.9% malnutrition prevalence, but low BMI was the sole indispensable phenotypic driver for only 233 individuals (5.2%). The GLIM-adapted framework maintained a highly comparable prevalence of 17.9% (n = 4,038). Thus, low BMI exhibits low diagnostic sensitivity in modern U.S. cohorts. Excluding BMI successfully bypasses circularity, unmasks qualitative muscle wasting under excess body weight, and establishes a robust, weight-independent standard for clinical and epidemiological research.

## 1. Introduction

The contemporary global nutritional landscape, particularly within Western societies, is defined by a complex “double burden” of malnutrition [1, 2]. Excessive caloric intake and obesity frequently coexist with severe qualitative nutrient deficiencies and progressive lean tissue depletion, a phenomenon widely referred to as “hidden hunger” or silent malnutrition [1]. In populations with a high prevalence of overweight and obesity, standard weight-based anthropometric measurements often fail as diagnostic indicators [3]. Adipose tissue accumulation masks underlying muscle wasting (sarcopenia) and micronutrient voids, thereby delaying clinical recognition and targeted nutritional support [3].

To standardize the identification of nutritional depletion, the Global Leadership Initiative on Malnutrition (GLIM) proposed a consensual framework requiring at least one phenotypic criterion and one etiologic criterion [4]. While low BMI remains a core phenotypic component of the standard GLIM criteria, its application in populations with high rates of obesity presents a critical methodological challenge [5]. Specifically, utilizing BMI simultaneously as a diagnostic component and as a stratification variable for subgroup analyses introduces a “circularity bias” that artificially distorts risk estimates [5]. To overcome this limitation and unmask qualitative malnutrition across the weight spectrum, clinical investigators have increasingly advocated for “GLIM-adapted” diagnostic models [6, 7]. These frameworks prioritize objective, instrument-based assessments of body composition—most notably dual-energy X-ray absorptiometry (DXA)—to evaluate muscle mass while intentionally excluding BMI from the diagnostic algorithm [7].

A recent analysis on the National Health and Nutrition Examination Survey (NHANES) database found that the standard GLIM framework applied identified a malnutrition prevalence of around 19%, corresponding to over 4,400 malnourished individuals within the adult cohort [8]. The primary objective of this study is to validate a weight-independent “GLIM-adapted” framework using a massive, nationally representative cohort of U.S. adults. We aim to demonstrate that removing BMI from the phenotypic criteria successfully bypasses circularity bias while preserving diagnostic sensitivity, thereby establishing a new methodological benchmark for epidemiological research in populations with a high prevalence of overweight and obesity.

## 2. Materials and Methods

### 2.1 Study Design and Population

We conducted a cross-sectional analysis utilizing data from 11 cycles of the NHANES spanning from 1999 to 2020. NHANES employs a complex, multistage, probability-based sampling design representative of the non-institutionalized U.S. civilian population. From an initial pool of 119,555 participants, we applied sequential exclusion criteria. The population was restricted to adults aged ≥ 20 years to align with technical standards for body composition and because adult malnutrition criteria are not validated for pediatric populations undergoing active growth [4, 5]. We excluded pregnant women to avoid gestational weight gain and fluid shifts confounding BMI and DXA-derived muscle mass [5]. After excluding subjects with missing data regarding dietary intake, inflammatory markers, or DXA-derived body composition parameters, the final analytical cohort comprised 23,363 participants.

### 2.2 Standard vs. “GLIM-Adapted” Diagnostic Operationalization

Nutritional status was evaluated and compared using two distinct diagnostic approaches:

1. **Standard GLIM Framework:** Malnutrition was diagnosed in the presence of at least one phenotypic criterion and at least one etiologic criterion [4].
2. **GLIM-Adapted Framework:** Malnutrition was diagnosed by completely excluding low BMI from the phenotypic criteria. Diagnosis required at least one of the remaining phenotypic criteria and at least one etiologic criterion [7].

Phenotypic criteria were operationalized as follows:

– **Involuntary Weight Loss:** Defined as a documented, involuntary weight loss of >10% beyond 6 months [4, 5].
– **Low BMI (Standard only):** Evaluated using age-differentiated thresholds (<20 kg/m^2^ for individuals aged <70 years; <22 kg/m^2^ for those aged ≥70 years) [4, 5].
– **Reduced Muscle Mass:** Measured objectively via DXA (the clinical reference standard for body composition) and operationalized using the Appendicular Lean Mass Index (ALMI), with established sex-specific cutoffs of <7.0 kg/m^2^ for men and <5.5 kg/m^2^ for women [4, 9].

And, etiologic criteria were operationalized as follows:

– **Reduced Food Intake or Assimilation:** Operationalized as a total daily energy intake <25 kcal/kg of body weight, indicating an intake insufficient to cover baseline metabolic demands [8, 10].
– **Inflammation or Disease Burden:** Identified biochemically by a serum C-reactive protein (CRP) level >5 mg/L, serving as a proxy for systemic pro-inflammatory catabolic states [5].

## 3. Results and Rationale for BMI Exclusion

The GLIM-adapted and standard GLIM framework identified, respectively, 4,038 and 4,425 malnourished individuals within the 23,363-adult cohort. The overlapping combinations of phenotypic and etiologic criteria are reported in Figure 1.

**Figure 1.**
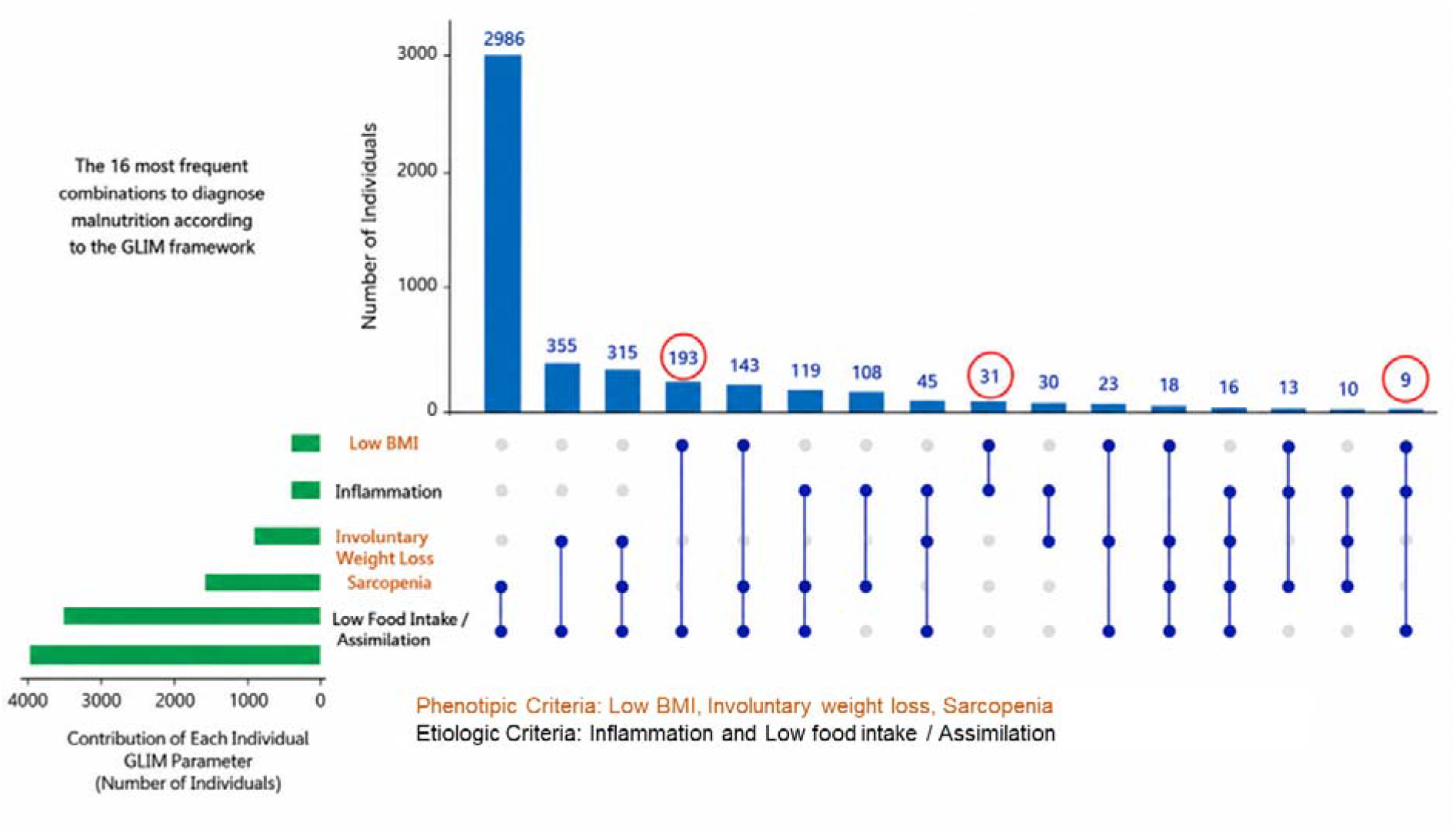
Overlapping combinations of GLIM criteria and the diagnostic contribution of BMI in U.S. adults (N = 23,363). The Euler-Venn diagram illustrates the distribution and intersections of phenotypic and etiologic criteria within the malnourished cohort (n = 4,425, representing a baseline prevalence of 18.9%)1. **Circled numbers** represent the specific subgroups (n = 233 in total) for whom a low Body Mass Index (BMI) was the sole and indispensable phenotypic criterion required to establish the diagnosis of malnutrition. In all other cases, malnutrition was classifiable via involuntary weight loss and/or objective DXA-derived muscle mass. Excluding these 233 individuals (representing only 5.2% of the malnourished cohort) prevents circularity bias in subsequent BMI-stratified analyses, yielding a weight-independent analytical cohort of 22,937 participants with a highly comparable malnutrition prevalence of 17.9%.

Involuntary weight loss and DXA-derived reduced muscle mass represented the primary phenotypic drivers of malnutrition, accounting for the vast majority of diagnoses. Conversely, low BMI emerged as the least sensitive phenotypic component in this U.S. cohort. Crucially, low BMI was identified as the sole indispensable phenotypic driver for only 233 subjects (5.2% of the malnourished group). For these 233 individuals, a diagnosis was driven only by the low BMI criterion because they did not meet the weight loss or reduced muscle mass thresholds. By removing these 233 individuals, the weight-independent GLIM-adapted framework classified 4,038 participants as malnourished, maintaining a comparable national prevalence of 17.9% (95% CI: 17.3%–18.3%) (Table 1). This empirical validation demonstrates that excluding BMI preserves 94.8% of the malnourished cohort while completely removing the risk of statistical circularity.

**Table 1.** Malnutrition Prevalence and Severity Across Frameworks (N=23,363)

| Diagnostic Status | Standard GLIM<br>n (%) | GLIM-Adapted<br>n (%) | Prevalence Difference<br>(95% CI) |
| --- | --- | --- | --- |
| Not-Malnourished | 18,938 (81.1) | 19,325 (82.2) | +1.1% (0.9% to 1.3%) |
| **Malnourished (Total)** | **4,425 (18.9)** | **4,038 (17.9)** | **-1.1% (-1.3% to -0.9%)** |
| Stage 1 (Moderate)<sup>†</sup> | 3,463 (78.3) | 3,076 (76.2) | — |
| Stage 2 (Severe)<sup>†</sup> | 962 (21.7) | 962 (23.8) | — |

**Abbreviation:** GLIM, Global Leadership Initiative on Malnutrition.
Severe (Stage 2) malnutrition under the GLIM framework is diagnosed when a patient meets at least one aetiologic criterion and a severe threshold in at least one phenotypic criterion.
<sup>†</sup> Percentages for Stage 1 and Stage 2 are calculated within the malnourished cohort.
Prevalence difference is statistically significant ( $p < 0.001$ ).

## 4. Discussion

This study represents the first large-scale validation of a weight-independent GLIM-adapted framework in a representative general population. Our findings show that in a modern Western society—where over 70% of adults live with overweight or obesity [2]—simplistic weight-based indices like BMI fail to capture metabolic depletion. This misalignment is further highlighted by the Dietary Guidelines for Americans 2025–2030 (released in 2026), which inverted the traditional food pyramid to restrict ultra-processed foods (which constitute 60% of U.S. caloric intake) and emphasize high-quality proteins (1.2 to 1.6 g/kg/day) to halt the obesity and sarcopenia epidemic [2].

Relying on BMI-dependent diagnostic criteria in epidemiologic research creates an artificial protective shield around heavier individuals with obesity, which are unlikely being classified as malnourished. By validating a weight-independent model, we proved that malnutrition is a qualitative tissue disorder, not a weight-deficit condition. Moving beyond BMI and adopting instrument-based body composition metrics (like DXA-ALMI) is a methodological imperative to unmask hidden hunger and prevent the silent progression of sarcopenic obesity in contemporary populations [3].

## Data Availability

The datasets analyzed during the current study are publicly available on the official National Health and Nutrition Examination Survey (NHANES) website hosted by the Centers for Disease Control and Prevention (CDC) and can be accessed at: https://www.cdc.gov/nchs/nhanes/index.htm.

https://www.cdc.gov/nchs/nhanes/index.html

## Data Availability Statement

The datasets analysed during the current study are publicly available from the National Health and Nutrition Examination Survey (NHANES), National Center for Health Statistics, Centers for Disease Control and Prevention.

## Author Contributions

F.O. conceived the study, interpreted the data, and drafted the manuscript. A.N. and L.P. contributed to data interpretation and critically revised the manuscript. All authors approved the final version.

